# The effect of antidepressants on severity of COVID-19 in hospitalized patients: A systematic review and meta-analysis

**DOI:** 10.1101/2022.04.11.22273709

**Authors:** Hosein Nakhaee, Reza Bayati, Mohammad Rahmanian, Amir Ghaffari Jolfayi, Moein Zangiabadian, Sakineh Rakhshanderou

**Affiliations:** Student Research Committee, School of Medicine, Shahid Beheshti University of Medical Sciences, Tehran, Iran; Department of Microbiology, School of Medicine, Shahid Beheshti University of Medical Sciences, Tehran, Iran; Environmental and Occupational Hazards Control Research Center, School of Public Health, Shahid Beheshti University of Medical Sciences, Tehran, Iran

**Keywords:** antidepressants, SSRI, COVID-19, SARS-CoV-2

## Abstract

**Introduction:** Clinical depression and the subsequent low immunity is a comorbidity that can act as a risk factor for severity of COVID-19 cases. Antidepressants such as SSRI and SNRI are associated with immune-modulatory effects, which dismiss inflammatory response and reduce lung tissue damage. The current systematic review and meta-analysis aims to evaluate the effect of antidepressant drugs on prognosis and severity of COVID-19 in hospitalized patients.

**Methods:** A systematic search was carried out in PubMed/Medline, EMBASE, and Scopus up to January 16, 2022. The following keywords were used: “COVID-19”, “SARS-CoV-2”, “2019-nCoV”, “SSRI”, “SNRI”, “TCA”, “MAOI”, and “Antidepressant”. The pooled risk ratio (RR) with 95% CI was assessed using a fixed or random-effect model. We considered P < 0.05 as statistically significant for publication bias. Data were analyzed by Comprehensive Meta-Analysis software, Version 2.0 (Biostat, Englewood, NJ).

**Results:** Twelve studies were included in our systematic review. Three of them were experimental with 1751, and nine of them were observational with 290,950 participants. Seven out of twelve articles revealed the effect of antidepressants on reducing severity of COVID-19. SSRI medications, including Fluvoxamine, Escitalopram, Fluoxetine, and Paroxetine and also among the SNRI drugs Venlafaxine are also reasonably associated with reduced risk of intubation or death. There were four studies showing no significant effect and one study showing the negative effect of antidepressants on prognosis of covid-19. The meta-analysis on clinical trials showed that fluvoxamine could significantly decrease the severity outcomes of COVID-19 (RR: 0.745; 95% CI: 0.580-0.956)

**Conclusions:** Most of the evidence supports that the use of antidepressant medications, mainly Fluvoxamine may decrease the severity and improve the outcome in hospitalizes patients with sars-cov-2. Some studies showed contradictory findings regarding the effects of antidepressants on severity of COVID-19. Further experimental studies should be conducted to clarify the effects of antidepressants on severity of COVID-19.

## Introduction

After over two years since the first case of the novel coronavirus was detected in December 2019 in Wuhan, China, 483 million infected cases and 6.13 million deaths have been reported worldwide up to January 21, 2022 (1). Covid19 is an acute respiratory disease, resulting in progressive respiratory failure, and eventually leading to death. Common symptoms include fever, dry cough, myalgia and fatigue. Severe cases develop dyspnea, hemoptysis, acute respiratory distress syndrome which can result in death. The severity or mortality is higher in older population, with comorbidities such as diabetes, hypertension, cardiovascular diseases and weaker immune functions (2). During the COVID-19 pandemic with strict lockdown measures, psychiatric patients have suffered episodes of anxiety, depression, and stress disorders on higher scales.(3) Clinical depression and the subsequent low immunity can also act as risk factors for severity of COVID-19 cases (4). Patients with clinical depression have lower immunity compared to those of healthy controls. A history of depression is associated with higher risk of infection and also the increased risk remains consistent over time (5). The suggested mechanism of action is fewer circulating CD4^+^ T-cells and reduction in natural killer cell cytotoxic responses of lymphocyte proliferation in elderly patients with clinical depression (6). Treatment of depression as an underlying morbidity could decrease the risk of clinical deterioration of Covid-19 patients. On the other hand, antidepressants such as SNRI and SSRI which are widely used in treatment of psychiatric patients are shown to reduce corona virus infection rates as patients who were receiving these medications were less likely to test positive for COVID-19 (7). Antidepressants are also associated with less severe cases as they significantly reduced the risk of intubation or death in some cohort studies (8, 9). Several mechanisms are suggested to justify this finding. Antidepressants could be associated with declined plasma levels of inflammatory cytokines, including IL-10, TNF-α, CCL-2, and IL-6, which are related to COVID-19 severity and mortality (10, 11). Also Some SSRI antidepressants, such as fluvoxamine which is a functional inhibitor of acid sphingomyelinase activity (FIASMA), may prevent the infection of epithelial cells with SARS-CoV-2 (12). In this study we aim to evaluate the effect of antidepressant drugs on prognosis and severity of COVID-19 in hospitalized patients.

## Method

This study was conducted and reported in accordance with the Preferred Reporting Items for Systematic Reviews and Meta-Analyses statement (13). The study was registered in the Systematic Review Registration: PROSPERO (pending registration ID: 313272).

### Search strategy

We searched Pubmed/Medline, Embase and Scopus for clinical studies reporting the effect of anti-depressants on reducing severity of hospitalized patients with covid-19, published up to January 16, 2022. We included clinical trials, cohort and case–control studies that were written in English. We used the following MeSH terms: “‘antidepressive agents’, ‘antidepressive agents, second generation’, ‘antidepressive agents, tricyclic’, ‘monoamine oxidase inhibitors’, ‘serotonin and noradrenaline reuptake inhibitors’, ‘serotonin uptake inhibitors’, ‘COVID-19’ and ‘SARS-CoV-2’” (Table S1&2&3). Keyword searches were done with combinations of the terms “SSRI”, “SNRI”, “TCA”, “MAOI”, “Antidepressant”, “2019 novel coronavirus” and “sars coronavirus 2”. Lists of references of selected articles and relevant review articles were hand-searched to identify further studies.

### Study Selection

The records found through database searching were merged, and the duplicates were removed using EndNote X8 (Thomson Reuters, Toronto, ON, Canada). Two reviewers independently screened the records by title/abstract and full text to exclude those unrelated to the study objectives. Any disagreements were resolved by the lead investigator. Included studies met the following criteria: (i) patients were diagnosed with COVID-19 based on the WHO criteria; (ii) patients were received anti-depressants; and (iii) severity predictors (hospitalization due to COVID-19 and/or ARDS and/or need to NIV or mechanical ventilation and/or ICU admission and/or death). Conference abstracts, editorials, reviews, study protocols, molecular or experimental studies on animal models and studies with focusing on infection risk were excluded.

### Data extraction

Two reviewers designed a data extraction form. These reviewers extracted data from all eligible studies, and differences were resolved by consensus. The following data were extracted: first author name; year of publication; study duration; type of study, country/ies where the research was conducted; demographics (i.e. age, sex); Detection test of COVID-19; anti-depressant type, dosage and frequency; Follow-up time; the definition of case and control; total number of controls and cases, severity indices, mechanism of action of anti-depressants against COVID-19 and Possibility of using anti-depressants in COVID-19 treatment.

### Quality assessment

Two blinded reviewers assessed the quality of the studies using three different assessment tools (checklists): two for observational studies (case controls and cohorts) and one for experimental studies (14). Items such as study population, measure of exposures, confounding factors, extent of outcomes, follow-up data, and statistical analysis were evaluated.

### Statistical Analysis

The pooled risk ratios (RRs) with 95% CI were assessed using random or fixed-effect models. The fixed-effects model was used because of the low estimated heterogeneity of the true effect sizes. The between-study heterogeneity was assessed by Cochran’s Q and the I2 statistic. Publication bias was evaluated statistically by using Egger’s and Begg’s tests as well as the funnel plot (p < 0.05 was considered indicative of statistically significant publication bias; funnel plot asymmetry also suggests bias) (15). All analyses were performed using “Comprehensive Meta-Analysis” software, Version 2.0 (Biostat, Englewood, NJ).

## Results

The selection process of articles is shown in Figure 1. Twelve articles were included and classified into the followings: six cohort studies (8, 16-20), three case-control studies (21-23) and three clinical trials (24-26). There were 872 cases and 879 controls in clinical trials, 15950 cases and 96638 controls in the cohort studies and 97,844 cases and 80518 controls in the case–control studies with the total population of 292701 in the whole studies (114666 cases and 178035 controls). Four studies were conducted in USA and other studies were designed in France, Turkey, Brazil, Hungary, Israel, Croatia, Scotland and Italy. The duration of studies, detection test of COVID-19 and other study characteristics are shown in table 1.

**Table 1.** Study characteristics

| First author | Study design | Publication year | country | Study duration | detection test of COVID-19 |
| --- | --- | --- | --- | --- | --- |
| Lenze et al. (25) | Randomized Clinical Trial | 2020 | USA | April 10, 2020, to August 5, 2020 | PCR |
| Reis et al. (26) | Randomized Clinical Trial | 2021 | Brazil | June 2, 2020 to Aug 5, 2021 | RT-PCR |
| Calusic et al. (24) | cohort trial | 2021 | Croatia | April and May 2021, | PCR |
| Israel et al. (21) | case-control | 2021 | Israel | November 30, 2020 to December 31, 2020. | RT-PCR |
| McKeigue et al. (22) | case-control | 2021 | Scotland | June 6, 2020 to June 14, 2020 | N/R |
| Nemeth et al. (23) | case-control | 2021 | Hungary | March 17, 2021 to April 22, 2021 | antigen or polymerase chain reaction test |
| Rauchman et al. (19) | retrospective cohort | 2021 | USA | March 2020 to March 2021 | N/R |
| Hoertel et al. (8) | retrospective cohort | 2021 | France | January 24, 2020 to April 1, 2020 | RT-PCR |
| FEI et al. (17) | Prospective Cohort | 2021 | Italy | N/R | N/R |
| Oskotsky et al. (18) | retrospective cohort | 2020 | USA | January to September 2020 | laboratory test for SARS-CoV-2 (nucleic acid amplification tests and immunoassays) and/or by ICD-10 (for COVID-19 confirmed by laboratory testing) |
| Seftel et al. (20) | Prospective Cohort | 2021 | USA | November–December 2020 | PCR |
| Bora et al. (16) | retrospective cohort | 2021 | Turkey | October 1, 2020 to January 1, 2021 | the ICD-10 classification confirmed by the test results |

**Figure1.**
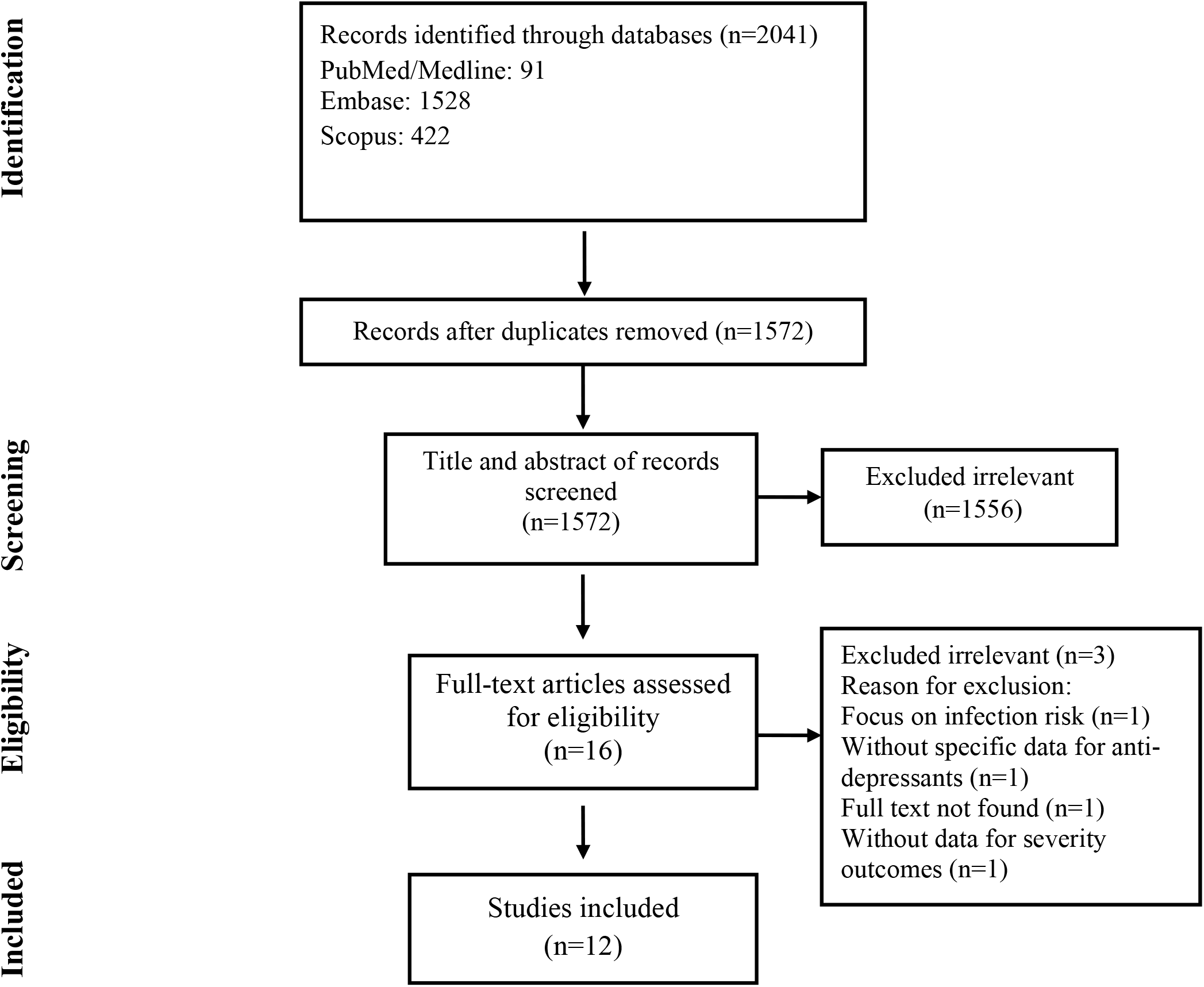
Flow chart of study selection for inclusion in the systematic review and meta-analysis.

### Quality of the included studies

The checklists for observational studies (14) showed that the included observational studies had a low risk of bias except Bora et al. study (16). (Table 2 & 3) In contrast, the checklist for experimental studies (14) showed that the included experimental studies had a high risk of bias for randomization, group concealment and participants, treatment delivers and outcome assessors blinding except Lenze et al. study (25). (Table 4)

**Table 2.** Quality assessment of cohort studies

| author | 1 | 2 | 3 | 4 | 5 | 6 | 7 | 8 | 9 | 10 | 11 |
| --- | --- | --- | --- | --- | --- | --- | --- | --- | --- | --- | --- |
| **Rauchman** | yes | yes | yes | no | no | yes | yes | yes | yes | yes | yes |
| **Hoertel** | yes | yes | yes | no | yes | yes | yes | yes | yes | no | yes |
| **FEI** | yes | yes | yes | no | no | yes | yes | yes | yes | yes | yes |
| **Oskotsky** | yes | yes | yes | no | yes | unclear | yes | yes | yes | yes | yes |
| **Seftel** | yes | yes | yes | no | yes | yes | yes | no | yes | yes | yes |
| **Bora** | yes | yes | yes | no | no | yes | yes | unclear | unclear | unclear | yes |

**Table 3.** Quality assessment of case-control studies

| author | 1 | 2 | 3 | 4 | 5 | 6 | 7 | 8 | 9 | 10 |
| --- | --- | --- | --- | --- | --- | --- | --- | --- | --- | --- |
| **Israel (Cohort1)** | yes | yes | yes | yes | yes | yes | yes | yes | yes | yes |
| **Israel (Cohort2)** | yes | yes | yes | yes | yes | yes | yes | yes | yes | yes |
| **McKeigue** | yes | yes | yes | yes | yes | yes | yes | yes | yes | yes |
| **Nemeth** | yes | yes | yes | yes | yes | no | no | yes | yes | yes |

1. Were the groups comparable other than the presence of disease in cases or the absence of disease in controls? 2. Were cases and controls matched appropriately? 3. Were the same criteria used for identification of cases and controls? 4. Was exposure measured in a standard, valid and reliable way? 5. Was exposure measured in the same way for cases and controls? 6. Were confounding factors identified? 7. Were strategies to deal with confounding factors stated? 8. Were outcomes assessed in a standard, valid and reliable way for cases and controls? 9. Was the exposure period of interest long enough to be meaningful? 10. Was appropriate statistical analysis used?

**Table 4.** Quality assessment of the experimental studies

| author | 1 | 2 | 3 | 4 | 5 | 6 | 7 | 8 | 9 | 10 | 11 | 12 | 13 |
| --- | --- | --- | --- | --- | --- | --- | --- | --- | --- | --- | --- | --- | --- |
| **Lenze** | yes | yes | yes | Yes | yes | yes | yes | yes | yes | yes | yes | yes | yes |
| **Calusic** | unclear | unclear | yes | Unclear | unclear | unclear | yes | yes | yes | yes | yes | yes | yes |
| **Reis** | yes | yes | yes | Unclear | unclear | unclear | yes | yes | yes | yes | yes | yes | yes |

1. Was true randomization used for assignment of participants to treatment groups? 2. Was allocation to treatment groups concealed? 3. Were treatment groups similar at baseline? 4. Were participants blind to treatment assignment? 5. Were those delivering treatment blind to treatment assignment? 6. Were outcome assessors blind to treatment assignment? 7. Were treatment groups treated identically other than the intervention of interest? 8. Was follow-up complete, and, if not, were differences between groups in terms of their follow-up adequately described and analyzed? 9. Were participants analyzed in the groups to which they were randomized? 10. Were outcomes measured in the same way for treatment groups? 11. Were outcomes measured in a reliable way? 12. Was appropriate statistical analysis used? 13. Was the trial design appropriate and were any deviations from the standard randomized controlled trial design accounted for in the conduct and analysis of the trial?

### Patient Characteristics

According to nine studies (8, 16-18, 21-25), the mean age of total patients was 58.1 years old with 40 and 56.5 years old in case and control groups in eight studies, respectively (8, 17, 18, 20, 21, 23-25). 49.6 percent of patients were female according to eleven studies (8, 16-21, 23-26). This number was 52% in case groups and 49.3% in control groups according to nine studies (17-21, 23-26). Cases and controls were matched with age, sex, comorbidities, smoking status, BMI category and etc. The definition of case and control groups and severity outcomes are shown in table 5.

**Table 5.**
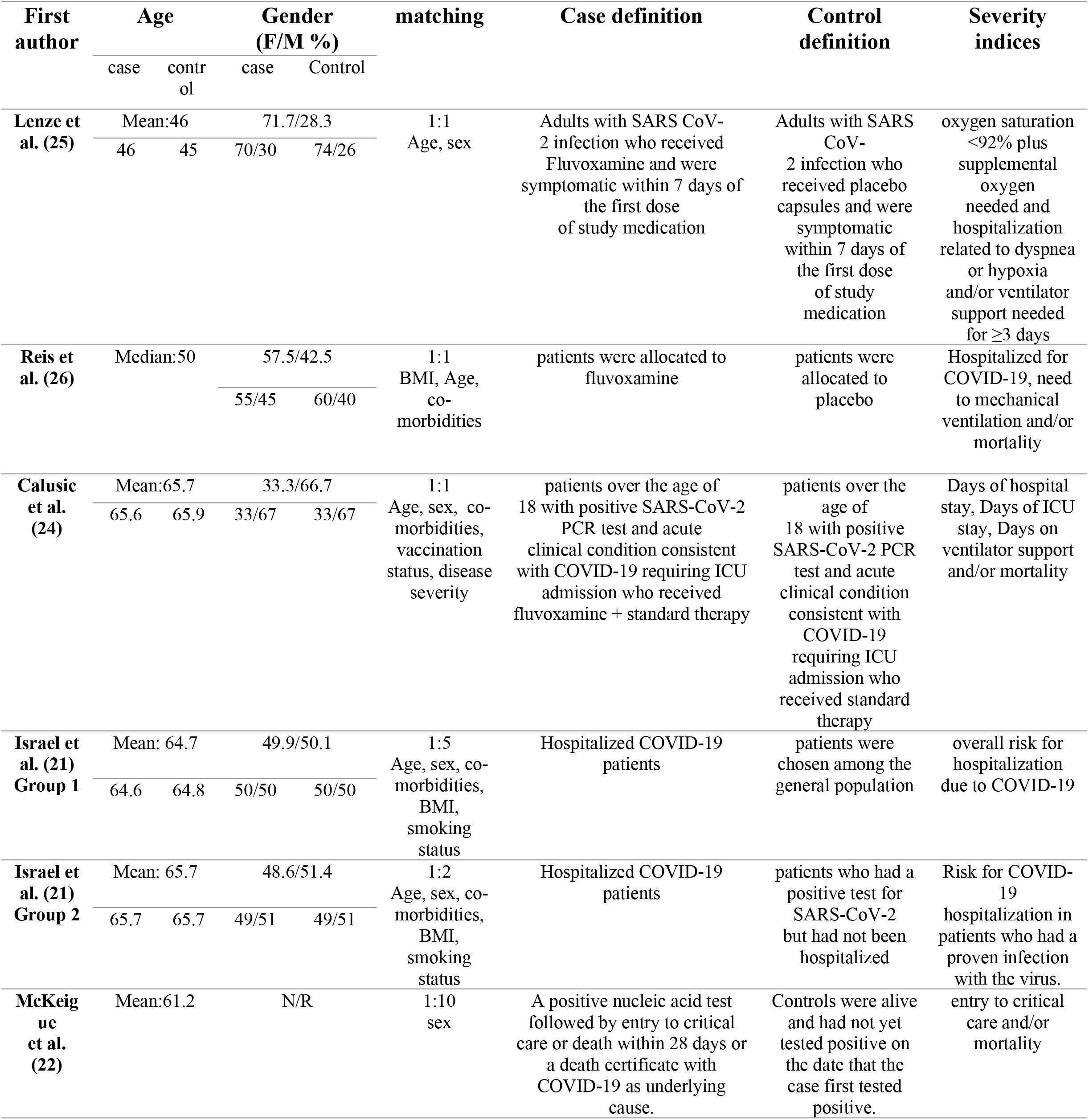
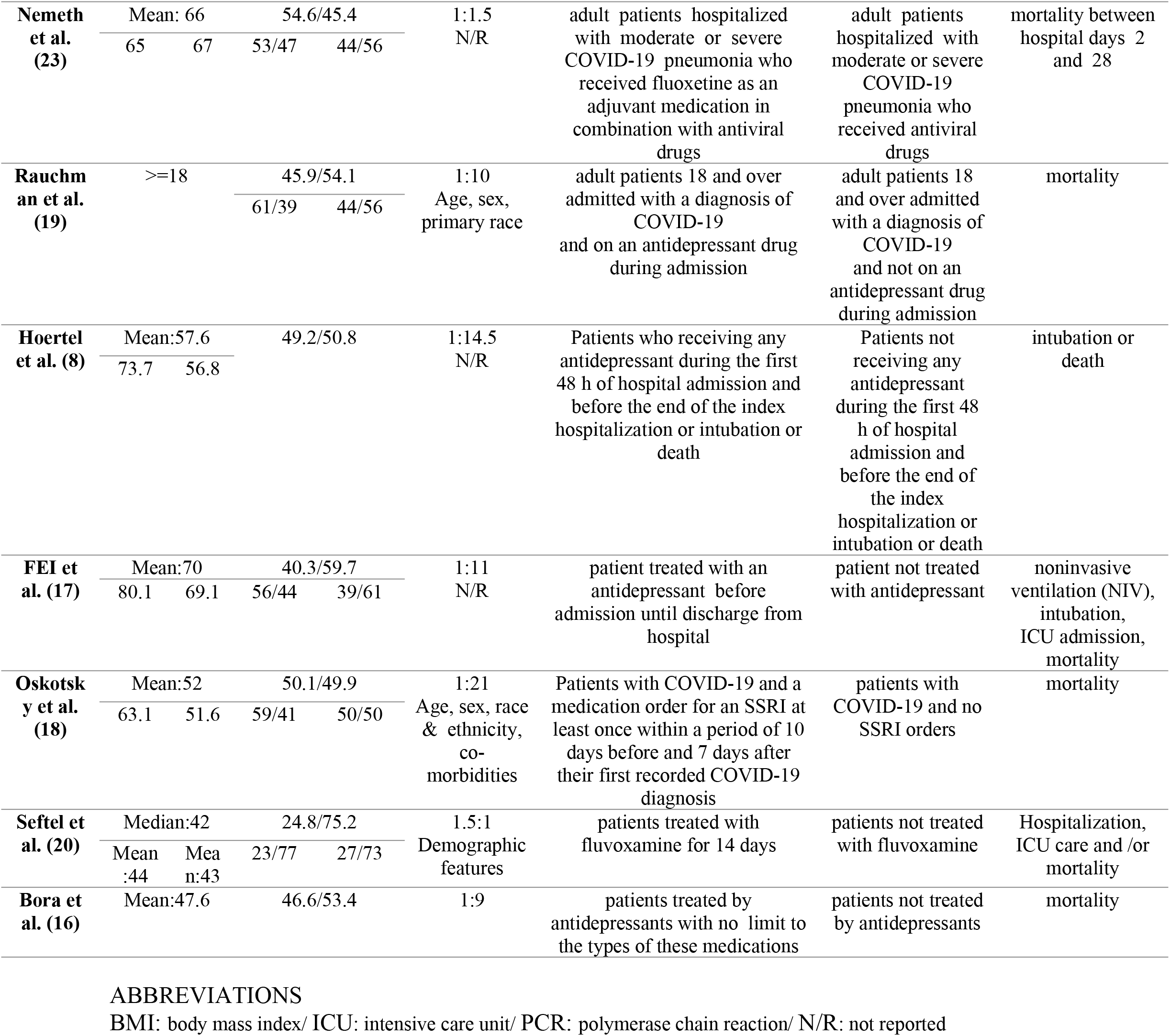
Patient characteristics

### Interventions and exposures Characteristics

Seven studies (18, 20, 21, 23-26) only used or surveyed SSRIs as anti-depressant in case groups (fluvoxamine in five studies (18, 20, 24-26), fluoxetine in two studies (18, 23) and Escitalopram in one study (21)). In remain five studies (8, 16, 17, 19, 22) different types of SSRIs, SNRIs and TCAs were used or surveyed. Anti-depressant type, dosage, frequency and follow-up time are shown in table 6.

**Table 6.**
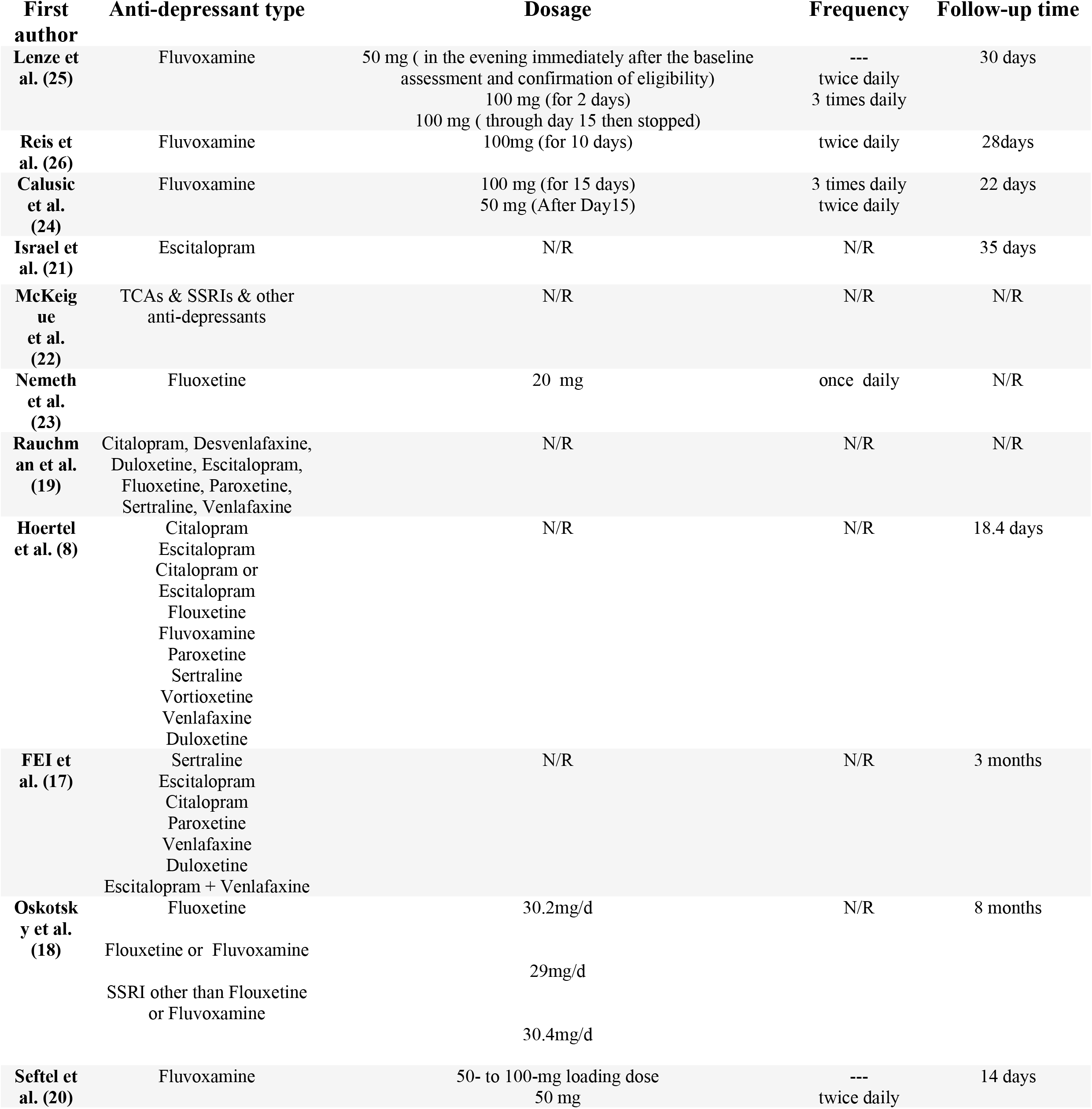
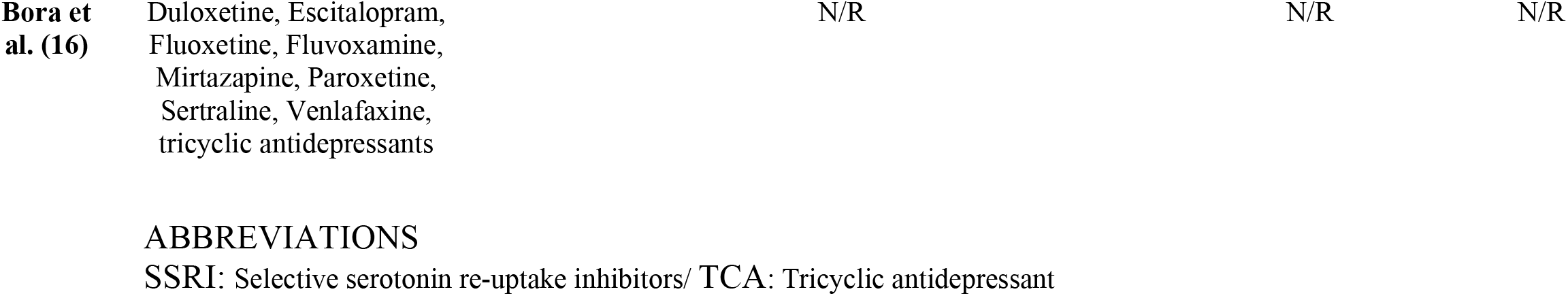
Interventions and exposures Characteristics

### Effect of anti-depressants on severity outcomes of COVID-19

After adjustment, seven studies (8, 16, 18, 20, 21, 23, 25) showed significant association between anti-depressant use and reducing severity outcomes of COVID-19. On the other hand, five studies (17, 19, 22, 24, 26) presented that either there are not any significant effect for anti-depressants to reduce disease severity or anti-depressants are associated with severe COVID-19 in hospitalized patients. In the study that was conducted by Bora et al. (16) anti-depressants were associated with reducing mortality, regardless of the anti-depressant type. Hoertel et al. (8) suggested that antidepressant use (HR:0.56; 95% CI: 0.43–0.73) is significantly and substantially associated with reduced risk of intubation or death, independently of patient characteristics, clinical and biological markers of disease severity, and other psychotropic medications. They found that SSRI (HR:0.51; 95% CI:0.316–0.72) and non-SSRI (HR:0.65; 95% CI, 0.45–0.93) antidepressants, and specifically the SSRIs escitalopram, fluoxetine, and paroxetine, the SNRI venlafaxine and the α2-antagonist antidepressants mirtazapine are significantly associated with reduced risk of intubation or death. Escitalopram is also effective in reducing severity according to Israel et al. (21) study. Fluoxetine use was associated with an important (70%) decrease of mortality (OR:0.33; 95% CI:0.16–0.68) and threefold survival in fluoxetine group according to Nemeth et al. (23) study. Also another study by Oskotsky et al. (18) showed similar result that fluoxetine could reduce risk of mortality (RR:0.72; 95% CI:0.54-0.97). On the other hand, Rauchman et al. (19) mentioned that prior use of SSRIs or SNRIs did not reduce mortality (OR:0.96; 95% CI:0.79-1.16). McKeigue et al. (22) proposed that TCAs (RR:1.1; 95% CI: 0.94-1.27) and SSRIs (RR:1.18; 95% CI:1.03-1.36) like other anti-depressants (RR: 1.76; 95% CI:1.5-2.07) increase the risk of severe COVID-19 because of their anti-cholinergic effect that is likely to increase risk of pneumonia. However, Fei et al. (17) represented that mortality rate, respiratory failure related to pneumonia and renal failure as a comorbidity within the anti-depressant treated subgroup is not lower than in the other patient of the sample but suggested that anti-depressant treated patient subgroup show high mean age and high size of medical comorbidity. Although, it is shown in this study that ARDS is significantly lower for the anti-depressant treated subgroup so mild significant lower employment of protease inhibitors and endotracheal intubation is needed in this subgroup because of lower level of IL-6 in anti-depressant treated patient that resulted from inhibition function of acid sphingomyelinase induced by antidepressants.

### Specific effect of fluvoxamine on severity outcomes of COVID-19

Four studies have discussed about the relation of using fluvoxamine and COVID-19 severity. One cohort study (20) and three clinical trials (24-26). Lenze et al. (25) represented that patients treated with fluvoxamine, compared with placebo, had a lower likelihood of clinical deterioration over 15 days (absolute difference:8.7% 95% CI:1.8%-16.4%). Also serious adverse events in fluvoxamine group were less than placebo group (1.3% vs 6.9%). They suggested that this effect is because of the influence of fluvoxamine on the S1R-IRE1 pathway and anti-inflammatory (Cytokine reduction) actions resulting from S1R activation. Also according to Seftel et al. (20) study the incidence of subsequent hospitalization was lower in fluvoxamine group (0% vs 12.5%) and elevated respiratory rates were improved faster by day 7 in this group so fluvoxamine seems to be promising as early treatment for COVID-19 to prevent clinical deterioration requiring hospitalization and to prevent possible long haul symptoms persisting beyond 2 weeks. On the other hand, Reis et al. (26) suggested that There were no significant differences between fluvoxamine and placebo for viral clearance at day 7 (RR:0·67; 95% CI:0·42–1·06), hospitalizations due to COVID-19 (RR:0·77; 95% CI:0·55–1·05), time to hospitalization (RR:0·79; 95% CI:0·58–1·06), mechanical ventilation (RR:0·77; CI:0·45–1·30), time on mechanical ventilation (RR:1·03; 95% CI: 0·64–1·67), death (RR:0·69; 95% CI:0·36– 1·27) and time to death (RR:0·80; 95%CI:0·43–1·51). However, according to Calusic et al. (24) study, there was no statistically significant differences between groups were observed regarding the number of days on ventilator support, duration of ICU or total hospital stay. But overall mortality was lower in the fluvoxamine group (HR: 0.58; 95% CI: 0.36–0.94). Although only mortality in women were significantly reduced in fluvoxamine group (HR: 0.40; 95% CI: 0.16– 0.99) and there were no significant differences in men mortality (HR: 0.69; 95% CI: 0.39–1.24).

### Statistical focus on fluvoxamine

The meta-analysis on clinical trials showed that fluvoxamine could significantly decrease the severity outcomes of COVID-19 (RR: 0.745; 95% CI: 0.580-0.956) (figure 2). There was no evidence of publication bias (p > 0.05) (figure 3).

**Figure 2.**
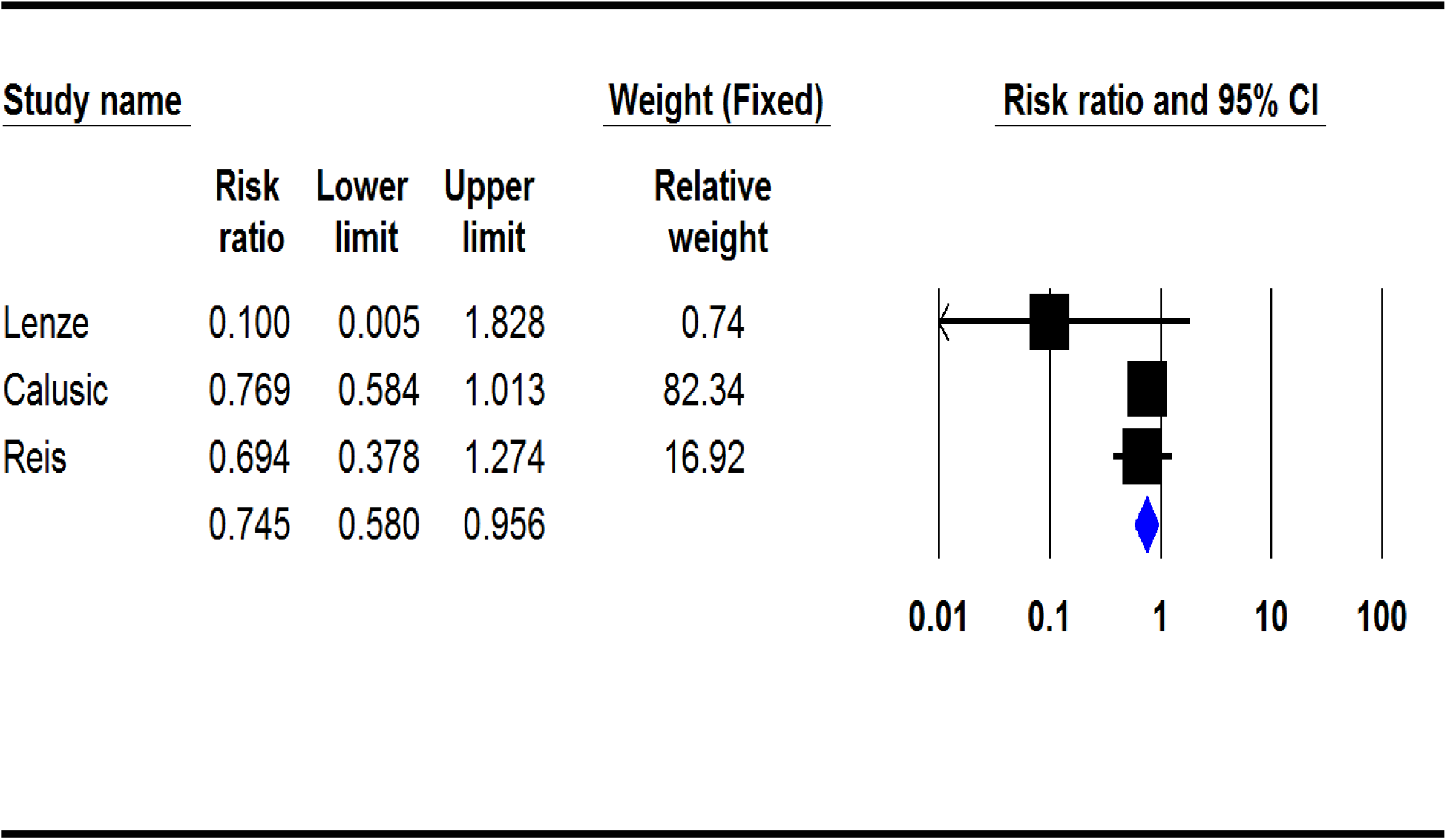
Pooled RR for clinical trials

**Figure 3.**
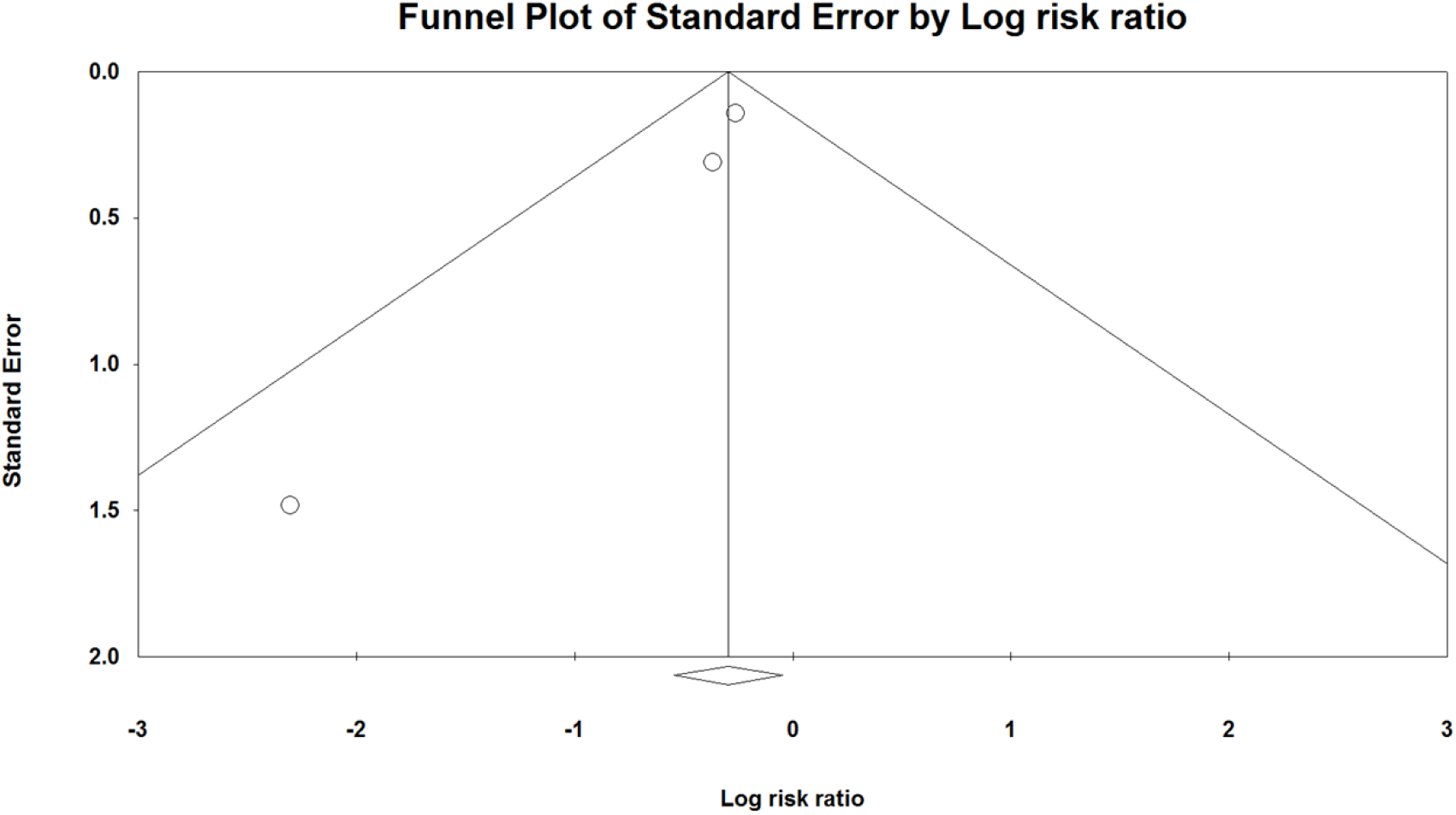
the funnel plot of analysis

## Discussion

Our systematic review study included cohort, case-control, and clinical trials and showed contradictory findings regarding the effect of antidepressant drugs on severity and mortality in covid-19 patients. Although various subgroups of antidepressants mainly reduce the risk of ARDS and mortality in hospitalized covid-19 patients compared to control groups, some studies reported no difference between case and control groups. On the other hand, one study proposed that antidepressants such as SSRIs and TCAs increase the risk of severe COVID-19 due to their anticholinergic effect that is likely to cause pneumonia.

Our meta-analysis on clinical trials also demonstrated that fluvoxamine could significantly decrease the severity outcomes of COVID-19 and as Seftel et al. (20) study proposed that it can be a therapeutic choice to prevent clinical deterioration requiring hospitalization for COVID-19 patients.

It has been proven that patients with clinical depression have lower immunity and thus more vulnerability to infection than healthy controls. (5) The suggested mechanism of action is fewer circulating CD4+ T-cells and reduction in natural killer cell cytotoxic responses of lymphocyte proliferation in elderly patients with clinical depression. There is also an abnormal regulation of the hypothalamus–pituitary–adrenal axis, sympathetic–adrenal–medullary axis, and hypothalamic-pituitary ovarian axis in patients with clinical depression (6). On the other hand, severe cases of COVID-19 infection are associated with an increase in the production of anti-inflammatory cytokines such as IL-6, IL-8, IL-10, and proliferation of CD8+ cytotoxic T-cells derived from CD4+ T-cells, which act in neutralizing the virus and inflammation of the lung (27). Thus it is justified to consider clinical depression and the subsequent low immunity as risk factors for the severity of COVID-19 cases.

In covid-19 patients with clinical depression, anti-depression treatment could improve immune responses by neutralizing the comorbidity (28). Still, there are several other mechanisms through which antidepressants might lower the risk of severe outcomes.

### Inhibition of serotonin transporter

SSRI and SNRIs are shown to have anti-inflammatory effects in both preclinical and clinical studies.(11, 29, 30) this might be owing to serotonin transporter inhibition.(31) Serotonin is involved in anti-inflammatory cytokine production. (32) For example, it can increase the release of the IL-10, which is a potent cytokine with reputable anti-inflammatory assets (33) IL-10 also moderates the levels of TNF-α and IL-6(34). Which are both responsible for the inflammatory response in covid-19 infection. Serotonin also reduces the production of inflammatory cytokines such as TNF-α and interferon-gamma (IFN-γ) by human blood leucocytes (35, 36). High serotonin transporter expression in the lungs(37) brings up its possible function in lung inflammation. Serotonin inhibits IL-12 and TNF-α release in human alveolar macrophages, but it increases IL-10 production via 5-HT2 receptors (38). Therefore, SSRIs and SNRIs may influence COVID-19 patients’ lung function. Considering that other antidepressants with the ability to block serotonin transporters did not show similar beneficial effects for COVID-19 patients, it is unlikely that serotonin transporter inhibition may play a significant role in SSRIs’ beneficial effects for COVID-19 patients. However, the anti-inflammatory effects of serotonin transporter inhibition may be a factor for the beneficial effects of SSRIs such as fluoxetine and fluvoxamine (31).

### Acid sphingomyelinase (ASM)

Many antidepressants, such as fluoxetine, fluvoxamine, sertraline, paroxetine, and amitriptyline, directly inhibit acid sphingomyelinase activity (11, 39, 40)which is necessary for epithelial cells to uptake SARS-CoV-2 (41). This could explain the findings of Hoertel et al. study (8), although considering that there are other drugs such as chlorpromazine with ASM inhibition activity that didn’t show the same protective effects on mortality in COVID-19 patients (42), this mechanism is unlikely a major factor for said effect.

### Sigma-1 receptor

sigma-1 receptor, an endoplasmic reticulum (ER) protein, is a cellular factor mediating the early steps of viral RNA replication, thus essential for virus replication at the early stage of infection (43, 44). A recent study showed that several compounds for sigma-1 and sigma-2 receptors are active in protein-protein interactions between SARS-CoV-2 and human proteins and could be promising inhibitors for SARS-CoV-2 replication (45).

On the other hand, animal studies indicated that Some SSRIs, such as fluvoxamine, sertraline, fluoxetine, and citalopram, have a high to moderate affinity for sigma-1 receptors in the rat brain. (45). Binding of Sigma-1 receptor agonists (i.e., fluvoxamine and fluoxetine) can result in sigma-1 receptor chaperone activity in the cells (46, 47). Another study demonstrated that binding of fluoxetine to the sigma-1 receptor in the endoplasmic reticulum decreases cytokine activity and enhances survival in preclinical models of sepsis and inflammation (48). Given the crucial role of the chaperone activity by sigma-1 receptor agonist in SARS-CoV-2 replication, it is possible that the SSRIs (i.e., fluvoxamine, fluoxetine, escitalopram) with sigma-1 receptor agonisms could be COVID-19 prophylactic drugs (8).

Another mechanism proposed to explain the anti-inflammatory effects of SSRIs is increasing melatonin levels due to cytochrome P450 enzyme CYP1A2 inhibition by fluvoxamine (49, 50). Melatonin which is naturally synthesized in the pineal gland and immune cells from the amino acid tryptophan, has anti-inflammatory, immunomodulatory, and antioxidant effects and is a therapeutic candidate to treat covid-19 (51).

Among the articles indicating no significant effect for antidepressants, Fei et al. (17) and Calusic et al. (24) studies were limited by small sample size. Calusic et al. (24) and Reis et al. (26) studies couldn’t entirely rule out selection bias. Fei et al. also stated that control group showed higher mean age and high size of medical comorbidities. Patients with antidepressant therapy have worse health conditions and immune responses than general population. This issue can have an impact on prognosis of covid-19 (17).

Another important clinical issue is to either continue or discontinue SSRIs when a COVID-19 patient is hospitalized or admitted to an ICU. There is evidence showing adverse ICU outcomes after discontinuation of SSRIs. This is probably due to increased agitation and need for sedation among these patients that can result in respiratory depression (52, 53). SSRI use may sometimes be contraindicated in ICU patients due to ECG changes and abnormal coagulation effects(54). A systematic review regarding the use of antidepressants in Critical Care, suggested that there may be excess morbidity in critically ill SSRI/SNRI users, but whether this is due to chronic effects, ongoing use, or drug withdrawal is unclear (53).

Reis et al (26) study suggested the modulatory effect of fluvoxamine on systemic inflammation as lower respiratory tract infections and hospital admissions were reported less frequently in patients in the fluvoxamine group compared to those in the placebo group.

Finally, efficacy of fluvoxamine in COVID-19 patients could be dependent on the timing of treatment, where increased effectiveness could be achieved if treatment is initiated earlier during SARSCoV-2 infection.

### Limitations

Most of the included articles in this study were not specific about antidepressant drugs and didn’t evaluate each drug separately. They mainly studied antidepressants in groups like SSRIs, SNRIs, and TCAs. We reviewed antidepressants but could only arrange a meta-analysis for fluvoxamine which was studied in three clinical trial which two of them had a high risk of bias. Our meta-analysis was limited to a small population of about 2000 persons. We also generally expressed our results about severity and outcome and couldn’t arrange a subgroup analysis for each outcome or sex group.

### Suggestions

As there is strong evidence of the link between antidepressant use and improving outcomes of covid-19, it seems legible to conduct more research on the subject, aiming to find more therapeutic options to treat covid-19. Future studies should focus on antidepressants separately and be more specific about the outcome in different patient groups. We also need to arrange more clinical trials with larger populations to confirm the efficacy of a candidate certainly. SSRIs such as Fluoxetine and Fluvoxamine are supported by stronger evidence and could be favorable options for future research programs.

## Conclusion

Among the studied experiment, seven studies showed anti-depressant ability to reduce the severity of covid-19, and five studies didn’t mention any significant effect on it. The covid-19 reducing effect of the mentioned medications can be concluded from the decline in the risk of intubation and death, clinical outcomes, and biological markers demonstrating the disease’s severity. Among the SSRI medication, the most common studied drug was fluvoxamine, which has a less subsequent hospitalization rate, less likelihood of 15-day-deterioration, and fewer adverse events than placebo, attributable to the S1R-IRE1 pathway and anti-inflammatory actions. Other SSRI medications, including Escitalopram, Fluoxetine, and Paroxetine and also among the SNRI drugs Venlafaxine are also reasonably associated with reduced risk of intubation or death. Still, they didn’t study as much as fluvoxamine. Although most of the evidence supports the effectiveness hypothesis of anti-depressant drugs, a single study proposed that some anti-depressants can intensify the severity of COVID-19 via anticholinergic effects that induce pneumonia. This issue should be investigated more precisely in future studies.

## Data Availability

All relevant data are within the manuscript and its Supporting Information files.

## DATA AVAILABILITY STATEMENT

The original contributions presented in the study are included in the article/supplementary material. Further inquiries can be directed to the corresponding authors.

## AUTHOR CONTRIBUTIONS

MZ, HN: designed the study. MZ, RB, MR, and AG: performed the search, study selection, and data synthesis. MZ, HN: wrote the first draft of the manuscript. SR: revised the article. All authors contributed to the article and approved the submitted version.

## CONFILICT OF INTREST

The mentioned in this article, have no involvement in any organization or entity with any financial interest nor non-financial interest such as personal or professional relationships, affiliations, knowledge or beliefs) in the subject matter discussed in this article.

## ACKNOWLEDGMENTS

This study is related to the MPH project From the Department of Public Health, School of Public Health and Safety, Shahid Beheshti University of Medical Sciences, Tehran, Iran.

